# Relative vaccine effectiveness of a CoronaVac booster dose in preventing symptomatic SARS-CoV-2 infection among healthcare workers during an Omicron period in Azerbaijan, January–August 2022

**DOI:** 10.1101/2023.10.20.23297300

**Authors:** Iris Finci, Madelyn Yiseth Rojas Castro, Nabil Seyidov, Samir Mehdiyev, C. Jason McKnight, Barbara Mühlemann, Christian Drosten, Lindsey Duca, Richard Pebody, Esther Kissling, Mark A. Katz, Gahraman Hagverdiyev

**Affiliations:** World Health Organization Regional Office for Europe, Copenhagen, Denmark; Epiconcept, Paris, France; Public Health and Reforms Center, Ministry of Health, Azerbaijan Republic; Institute of Virology, Charité–Universitätsmedizin Berlin, corporate member of Freie Universität Berlin, Humboldt-Universität zu Berlin, and Berlin Institute of Health, 10117 Berlin, Germany; German Centre for Infection Research (DZIF), partner site Charité, 10117 Berlin, Germany; United States Centers for Disease Control and Prevention, Atlanta, GA, United States of America

**Keywords:** Azerbaijan, CoronaVac, COVID-19, healthcare workers, SARS-CoV-2, booster, vaccination, vaccine effectiveness, BA.1, BA.2, Omicron

## Abstract

Among Azerbaijani healthcare workers (HCWs), compared to primary vaccine series, CoronaVac booster relative vaccine effectiveness was 60% (95% CI:25–79) and 79% (95% CI:44–92) against symptomatic and medically attended illness, respectively, during an Omicron BA.1/BA.2 period. Our results support timely CoronaVac booster uptake among Azerbaijani HCWs to reduce morbidity.

## Main text

COVID-19 vaccination has been critical in reducing COVID-19-related morbidity and mortality (1,2). Vaccinating healthcare workers (HCWs) against COVID-19 is vital for more resilient healthcare systems; HCWs face high SARS-CoV-2 exposure, regularly interact with vulnerable patients, and are essential for the continuous functioning of health services. In Azerbaijan, an upper-middle income country, HCWs have been prioritized for COVID-19 vaccination throughout the vaccination campaign. CoronaVac (CN02 strain, Sinovac, Beijing) has mainly been used. While inactivated vaccines like CoronaVac have been widely used in low- and middle-income countries, including those in the WHO European Region, few studies have evaluated their effectiveness, particularly during the period characterised by circulation of Omicron variants BA.1, BA.2 and BA.4/5 (hereafter ‘Omicron period’) (3–8).

Understanding COVID-19 vaccine effectiveness is essential for guiding vaccine policy. Additionally, having local data showing effectiveness of COVID-19 vaccines can help promote vaccine uptake. Using a prospective cohort of HCWs in Azerbaijan, we previously estimated that vaccine effectiveness (VE) of two-dose primary vaccine series (PVS) against symptomatic SARS-CoV-2 infection was 19% during a Delta-predominant period (9). In this study, we the same cohort to estimate relative VE (rVE) of a CoronaVac booster dose compared to PVS against symptomatic SARS-CoV-2 infection during an Omicron-predominant period.

### The study

This cohort study has been described previously (9,10). Briefly, in mid-2021, we enrolled HCWs from seven hospitals in Baku and collected information on sociodemographics, occupational exposure, and COVID-19 vaccination status. Participants completed weekly symptom questionnaires, those reporting any respiratory symptom were tested for SARS-CoV-2 by RT-PCR. We administered questionnaires to PCR-positive participants 30 days post-symptom onset to obtain information about their illness course. Data on vaccination and SARS-CoV-2 PCR testing results were validated using national databases.

We restricted the analysis to participants eligible for booster doses according to the Azerbaijan government: those who had received PVS at least five months (150 days) prior. During January 1 – August 31, 2022, we estimated the rVE of a first booster compared to a PVS series against two outcomes: i) symptomatic PCR-confirmed SARS-CoV-2 infection (COVID-19) and ii) medically attended COVID-19, defined as an outpatient visit because of COVID-19. Participants were considered fully vaccinated 14 days after receiving their PVS and 7 days after receiving their booster dose. We assessed the effect of time since vaccination on rVE against symptomatic infection over the time intervals: 7–89, 90–150 and ≥150 days following booster dose.

We evaluated rVE during the overall study period and separately for the period in which SARS-CoV-2 Omicron BA.1 and BA.2 variants were predominantly circulating (January 1–March 31, 2022), which we defined using whole genome sequencing data from study samples and publicly available data from GISAID (Figure S2) (9,11).

Relative VE was estimated as (1–adjusted hazard ratio)*100. Hazard ratios comparing vaccinated and unvaccinated participants were estimated using Cox proportional hazards models with vaccination as a time-varying exposure and a calendar timescale. Hospital location (urban or sub-urban) and previous SARS-CoV-2 infection were included in the model as fixed effects. Additional details about study methods are included in the supplemental material. The study was approved by the Azerbaijan and WHO Research Ethics Review Committees. All study participants provided written informed consent.

At the study start, there were 1367 participants (Figure S1). The median age was 48 (IQR: 39–57), and 1276 (93%) were female. Overall, 506 (37%) were nurses, 349 (26%) were physicians, and 512 (37%) were other professions. Overall, 375 (27%) had received PVS, 939 (69%) had received a booster, 30 (2%) received one vaccine dose, and 23 (2%) remained unvaccinated. Only participants who received CoronaVac vaccine were included in the analysis (Table 1). The median time from receiving the last vaccine dose until the start of follow-up was 66 days (IQR: 56–96) for booster dose and 150 days (IQR: 150–187) for PVS. At the study end, 1186 (87%) participants had received a CoronaVac booster dose (Table S1).

**Table 1.** Demographic and occupational characteristics of health care worker participants by vaccination status on the first day of the follow-up period (n=1367), Azerbaijan, 2022.

|  | All<br>Participants,<br>N=1367 | Unvaccinated,<br>N=23 | Partially<br>vaccinated (one<br>dose), N=30 | Vaccinated<br>with primary<br>vaccine series<br>(two doses),<br>N=375 | Vaccinated<br>with booster<br>dose (third<br>dose), N=939 |
| --- | --- | --- | --- | --- | --- |
| **Age, N=1367** |  |  |  |  |  |
| Median (IQR) | 48 (39-57) | 51 (37-62.5) | 43 (35-58.8) | 47 (38-56) | 49 (40-57) |
| **Age group, years, N=1367** |  |  |  |  |  |
| 20-29, n (%) | 73 (5) | 3 (13) | 1 (3) | 29 (8) | 40 (4) |
| 30-39, n (%) | 290 (21) | 6 (26) | 12 (40) | 84 (22) | 188 (20) |
| 40-49, n (%) | 371 (27) | 2 (9) | 4 (13) | 96 (26) | 269 (29) |
| 50-59, n (%) | 416 (30) | 4 (17) | 8 (27) | 110 (29) | 294 (31) |
| 60+, n (%) | 217 (16) | 8 (35) | 5 (17) | 56 (15) | 148 (16) |
| **Sex, N=1367** |  |  |  |  |  |
| M, n (%) | 91 (7) | 1 (4) | 4 (13) | 21 (6) | 65 (7) |
| F, n (%) | 1276 (93) | 22 (96) | 26 (87) | 354 (94) | 874 (93) |
| **Hospital, N=1367** |  |  |  |  |  |
| Hospital 7, n (%) | 269 (20) | 4 (17) | 2 (7) | 40 (11) | 223 (24) |
| Hospital 15, n (%) | 122 (9) | 0 (0) | 1 (3) | 8 (2) | 113 (12) |
| Hospital 18, n (%) | 204 (15) | 1 (4) | 3 (10) | 56 (15) | 144 (15) |
| Hospital 23, n (%) | 95 (7) | 3 (13) | 6 (20) | 29 (8) | 57 (6) |
| Hospital 24, n (%) | 191 (14) | 6 (26) | 5 (17) | 82 (22) | 98 (10) |
| Hospital 26, n (%) | 359 (26) | 2 (9) | 10 (33) | 107 (29) | 240 (26) |
| Hospital 29, n (%) | 127 (9) | 7 (30) | 3 (10) | 53 (14) | 64 (7) |
| **Hospital Location, N=1367** |  |  |  |  |  |
| Urban, n (%) | 517 (38) | 7 (30) | 9 (30) | 146 (39) | 355 (38) |
| Sub-urban, n (%) | 850 (62) | 16 (70) | 21 (70) | 229 (61) | 584 (62) |
| **Occupation/Role in hospital, N=1367** |  |  |  |  |  |
| Nurse or Midwife, n (%) | 506 (37) | 7 (30) | 11 (37) | 142 (38) | 346 (37) |
| Medical Doctor, n (%) | 349 (26) | 7 (30) | 8 (27) | 101 (27) | 233 (25) |
| Other, n (%) | 512 (37) | 9 (39) | 11 (37) | 132 (35) | 360 (38) |
| **BMI, N=1367** |  |  |  |  |  |
| Underweight or normal, n (%) | 382 (28) | 8 (35) | 6 (20) | 110 (29) | 258 (27) |
| Overweight, n (%) | 509 (37) | 9 (39) | 7 (23) | 129 (34) | 364 (39) |
| Obese, n (%) | 476 (35) | 6 (26) | 17 (57) | 136 (36) | 317 (34) |
| **Smoking, N=1367** |  |  |  |  |  |
| Currently smokes, n (%) | 38 (3) | 1 (4) | 1 (3) | 6 (2) | 30 (3) |
| Never smokes, n (%) | 1315 (96) | 22 (96) | 27 (90) | 368 (98) | 898 (96) |
| Smoked previously, n (%) | 14 (1) | 0 (0) | 2 (7) | 1 (<1) | 11 (1) |
| **Any chronic condition, N=1367** |  |  |  |  |  |
| No, n (%) | 820 (60) | 14 (61) | 14 (47) | 212 (57) | 580 (62) |
| Yes, n (%) | 547 (40) | 9 (39) | 16 (53) | 163 (43) | 359 (38) |
| **Number of chronic conditions, N=1367** |  |  |  |  |  |
| 0, n (%) | 820 (60) | 14 (61) | 14 (47) | 212 (57) | 580 (62) |
| 1, n (%) | 383 (28) | 4 (17) | 10 (33) | 104 (28) | 265 (28) |
| ≥2, n (%) | 164 (12) | 5 (22) | 6 (20) | 59 (16) | 94 (10) |
| **Number of household members, N=1280** |  |  |  |  |  |
| 1-3, n (%) | 500 (39) | 12 (55) | 10 (34) | 138 (39) | 340 (39) |
| 4-5, n (%) | 654 (51) | 6 (27) | 15 (52) | 174 (49) | 459 (52) |
| ≥6, n (%) | 126 (10) | 4 (18) | 4 (14) | 41 (12) | 77 (9) |
| **Hands on care, N=1367** |  |  |  |  |  |
| No, n (%) | 798 (58) | 14 (61) | 18 (60) | 200 (53) | 566 (60) |
| Yes, n (%) | 569 (42) | 9 (39) | 12 (40) | 175 (47) | 373 (40) |
| **Previous COVID-19 PCR confirmed infection, N=1367** |  |  |  |  |  |
| 0, n (%) | 1114 (81) | 19 (83) | 12 (40) | 233 (62) | 850 (91) |
| 1, n (%) | 253 (19) | 4 (17) | 18 (60) | 142 (38) | 89 (9) |
| **Delay between 2nd dose and start of follow-up (days), N=1314** |  |  |  |  |  |
| Median (IQR) | 305 (183-313) |  |  | 150 (150-187) | 309 (274-316) |
| **Delay between 3rd dose and start of follow-up (days), N=939** |  |  |  |  |  |
| Median (IQR) | 66 (56-92) |  |  |  | 66 (56-92) |
| **COVID-19 vaccination status and vaccine brand at study period start, N=1367** |  |  |  |  |  |
| Unvaccinated | 23 (2) | 23 (100) |  |  |  |
| CoronaVac - 2 doses, n (%) | 375 (27) |  |  | 375 (100) |  |
| CoronaVac - 3 doses, n (%) | 939 (69) |  |  |  | 939 (100) |
| CoronaVac - 1 dose, n (%) | 30 (2) |  | 30 (100) |  |  |

During the study period, 74 participants (5%) had PCR-confirmed COVID-19, of whom 53 (72%) had been vaccinated with a booster dose and 17 (23%) with PVS. Most cases [66 (89%)] occurred between January and March 2022, mirroring the trends in COVID-19 incidence in Azerbaijan (Figure 1). Among the participants with PCR-confirmed SARS-CoV-2, 24 (32%) sought outpatient medical care. There were no emergency room visits, hospitalisations, or deaths among participants.

**Figure 1.**
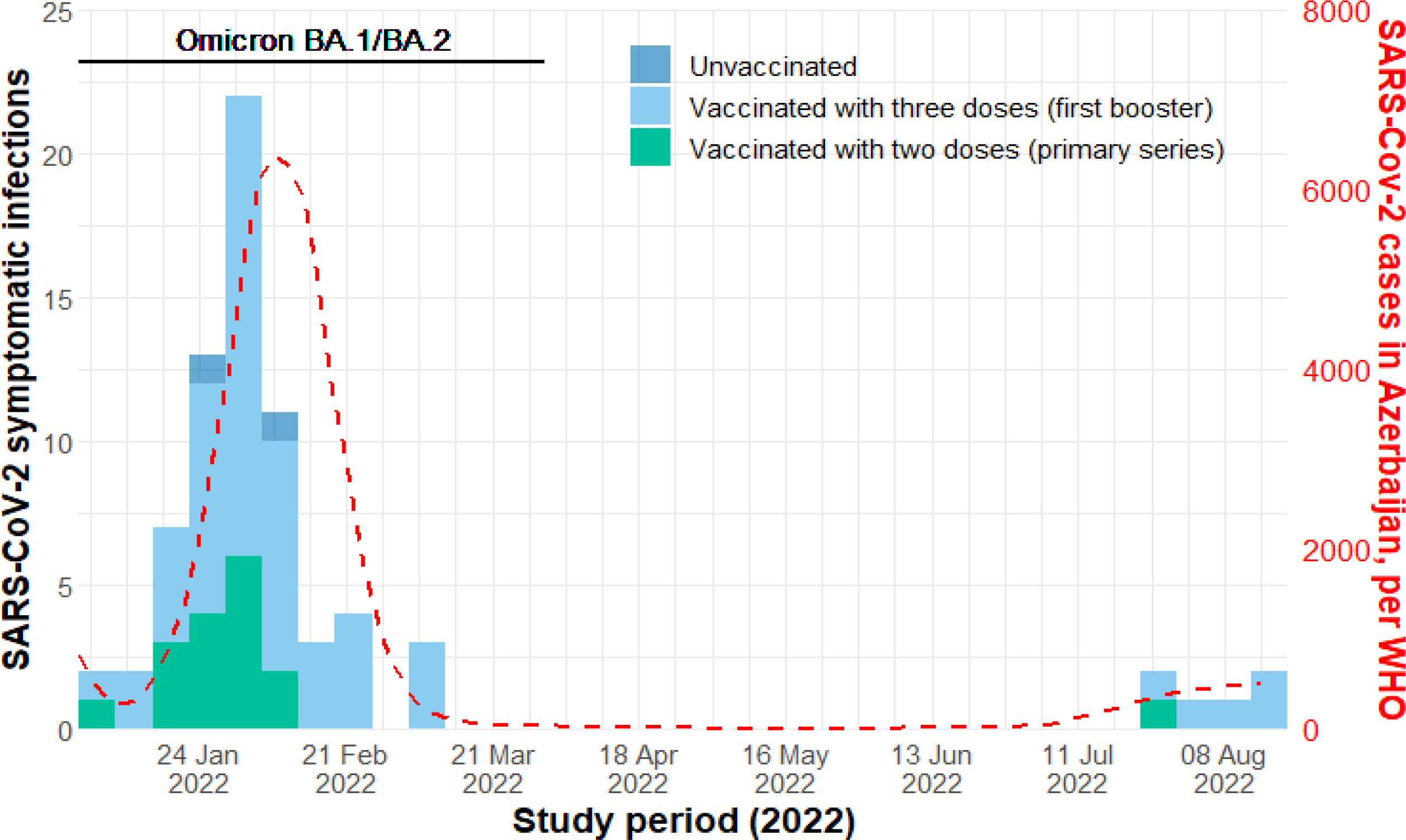
Number of symptomatic PCR-confirmed SARS-CoV-2 positive healthcare workers by vaccination status and symptom onset, and number of SARS-CoV-2 cases in Azerbaijan (14), January-August 2022.

For the whole analysis period (January–August 2022), the rVE was 59% (95% CI: 24–78) against symptomatic infection and 78% (95% CI: 41–92) against medically attended infection. During the Omicron BA.1/BA.2-predominant period (January–March 2022), rVE of a CoronaVac booster was 60% (95% CI: 25–79) against symptomatic infection and 79% (95% CI: 44–92) against medically attended COVID-19 (Figure 2, Table 2). Relative VE against symptomatic infection was 50% (95% CI: -3–75) 7-89 days post-booster, 67% (95% CI: 35–83) 90-149 days post-booster, and 48% (95% CI: -190–91) ≥150 days post-booster during the whole study period. (Figure 2, Table 2).

**Table 2.**
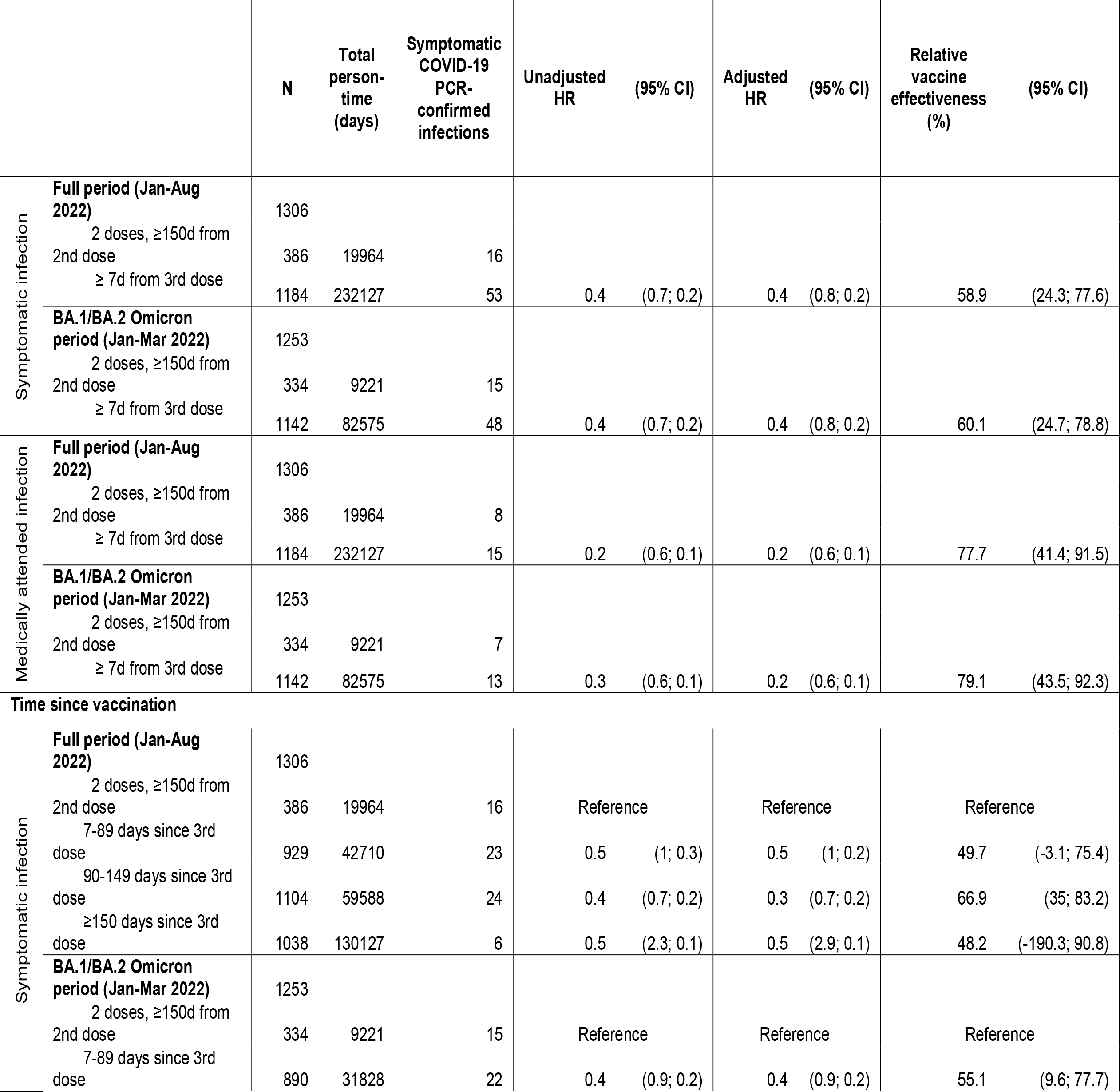
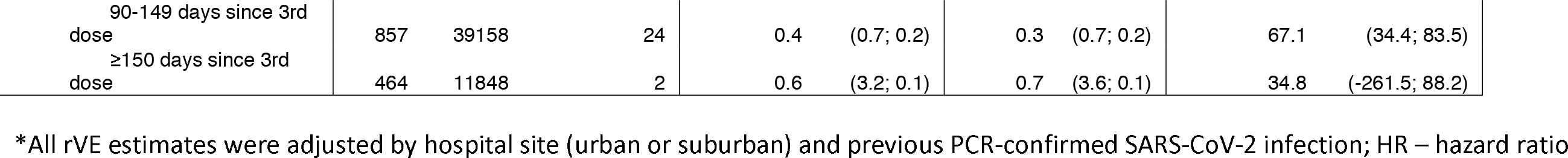
Relative COVID-19 vaccine effectiveness of CoronaVac booster dose compared to CoronaVac primary vaccine series against symptomatic and medically attended SARS-CoV-2 infection during a period of Omicron and relative vaccine effectiveness against symptomatic SARS-CoV-2 infection by time since receiving booster dose, Azerbaijan, January 1 – August 31, 2022

| Symptomatic SARS-CoV-2 infection by time since receiving booster dose, Alerba, Jan, January 1 – August 31, 2022 |  |  |  |  |  |  |  |  |  |  |
| --- | --- | --- | --- | --- | --- | --- | --- | --- | --- | --- |
|  |  | N | Total person-time (days) | Symptomatic COVID-19 PCR-confirmed infections | Unadjusted HR | (95% CI) | Adjusted HR | (95% CI) | Relative vaccine effectiveness (%) | (95% CI) |
| Symptomatic infection | Full period (Jan-Aug 2022) | 1306 |  |  |  |  |  |  |  |  |
|  | 2 doses, ≥150d from 2nd dose | 386 | 19964 | 16 |  |  |  |  |  |  |
|  | ≥ 7d from 3rd dose | 1184 | 232127 | 53 | 0.4 | (0.7; 0.2) | 0.4 | (0.8; 0.2) | 58.9 | (24.3; 77.6) |
|  | BA.1/BA.2 Omicron period (Jan-Mar 2022) | 1253 |  |  |  |  |  |  |  |  |
|  | 2 doses, ≥150d from 2nd dose | 334 | 9221 | 15 |  |  |  |  |  |  |
|  | ≥ 7d from 3rd dose | 1142 | 82575 | 48 | 0.4 | (0.7; 0.2) | 0.4 | (0.8; 0.2) | 60.1 | (24.7; 78.8) |
| Medically attended infection | Full period (Jan-Aug 2022) | 1306 |  |  |  |  |  |  |  |  |
|  | 2 doses, ≥150d from 2nd dose | 386 | 19964 | 8 |  |  |  |  |  |  |
|  | ≥ 7d from 3rd dose | 1184 | 232127 | 15 | 0.2 | (0.6; 0.1) | 0.2 | (0.6; 0.1) | 77.7 | (41.4; 91.5) |
|  | BA.1/BA.2 Omicron period (Jan-Mar 2022) | 1253 |  |  |  |  |  |  |  |  |
|  | 2 doses, ≥150d from 2nd dose | 334 | 9221 | 7 |  |  |  |  |  |  |
|  | ≥ 7d from 3rd dose | 1142 | 82575 | 13 | 0.3 | (0.6; 0.1) | 0.2 | (0.6; 0.1) | 79.1 | (43.5; 92.3) |
| Time since vaccination |  |  |  |  |  |  |  |  |  |  |
| Symptomatic infection | Full period (Jan-Aug 2022) | 1306 |  |  |  |  |  |  |  |  |
|  | 2 doses, ≥150d from 2nd dose | 386 | 19964 | 16 | Reference |  | Reference |  | Reference |  |
|  | 7-89 days since 3rd dose | 929 | 42710 | 23 | 0.5 | (1; 0.3) | 0.5 | (1; 0.2) | 49.7 | (-3.1; 75.4) |
|  | 90-149 days since 3rd dose | 1104 | 59588 | 24 | 0.4 | (0.7; 0.2) | 0.3 | (0.7; 0.2) | 66.9 | (35; 83.2) |
|  | ≥150 days since 3rd dose | 1038 | 130127 | 6 | 0.5 | (2.3; 0.1) | 0.5 | (2.9; 0.1) | 48.2 | (-190.3; 90.8) |
|  | BA.1/BA.2 Omicron period (Jan-Mar 2022) | 1253 |  |  |  |  |  |  |  |  |
|  | 2 doses, ≥150d from 2nd dose | 334 | 9221 | 15 | Reference |  | Reference |  | Reference |  |
|  | 7-89 days since 3rd dose | 890 | 31828 | 22 | 0.4 | (0.9; 0.2) | 0.4 | (0.9; 0.2) | 55.1 | (9.6; 77.7) |

|  |  |  |  |  |  |  |  |  |  |
| --- | --- | --- | --- | --- | --- | --- | --- | --- | --- |
| 90-149 days since 3rd<br>dose | 857 | 39158 | 24 | 0.4 | (0.7; 0.2) | 0.3 | (0.7; 0.2) | 67.1 | (34.4; 83.5) |
| ≥150 days since 3rd<br>dose | 464 | 11848 | 2 | 0.6 | (3.2; 0.1) | 0.7 | (3.6; 0.1) | 34.8 | (-261.5; 88.2) |

\*All rVE estimates were adjusted by hospital site (urban or suburban) and previous PCR-confirmed SARS-CoV-2 infection; HR – hazard ratio

**Figure 2.**
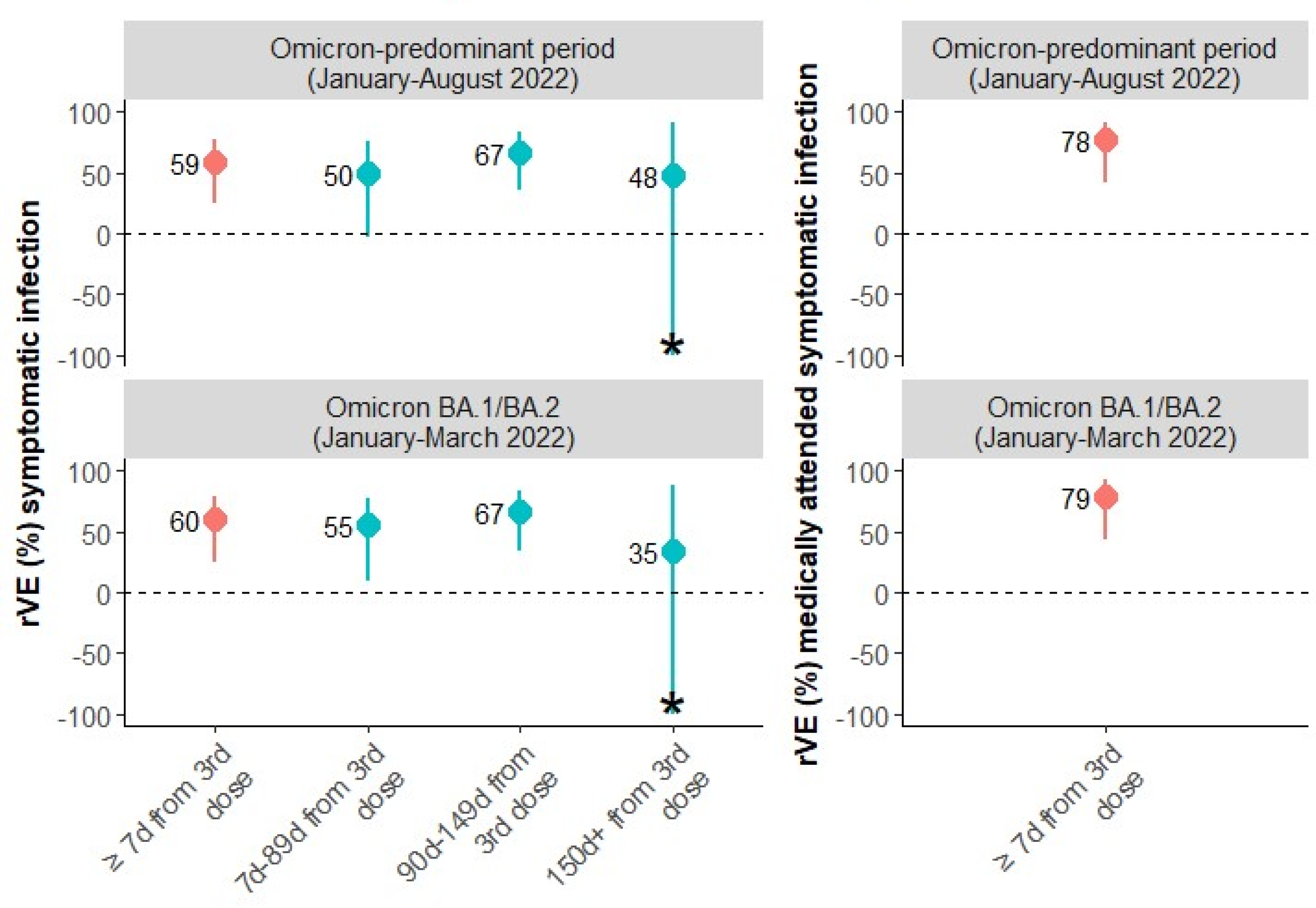
Relative vaccine effectiveness of a CoronaVac booster dose compared to CoronaVac primary series, Azerbaijan, 2022. Left panel – symptomatic SARS-CoV-2 infection, total rVE (red) and stratified by time since vaccination (blue); Right panel – medically attended SARS-CoV-2 infection. Star (*) indicates that the scale was cut off.

## Conclusions

Among HCWs in Azerbaijan, a CoronaVac booster dose offered additional protection against symptomatic infection and medically attended infection compared to CoronaVac PVS during an Omicron period. This protection, which lasted at least five months, occurred in a cohort that already had rates of prior infection approaching 70% in mid-2021 (9), highlighting the importance of the booster dose in reducing COVID-19 morbidity, even among HCWs with high rates of previous infection. The importance of the booster dose is relevant for the general population of Azerbaijan too, where COVID-19 PVS uptake is 47% and booster dose uptake is 34% (12).

Other studies have found variable rVE and absolute VE of a CoronaVac booster dose. A Brazilian study estimated a lower rVE of booster CoronaVac against symptomatic disease of 5% at 8-59 days after receipt; rVE subsequently declined to zero after 60 days in that study (5). Uncontrolled bias may have contributed to the low and negative estimates against symptomatic infection in that study (13). The same study estimated an rVE of booster CoronaVac against hospitalization of 47% during the same time period (5). In Hong Kong, in a mainly infection-naïve population, estimated absolute VE of CoronaVac booster against symptomatic BA.2 infection was 42% (3). In Brazil, in a population with higher seroprevalence, absolute VE of a CoronaVac booster dose was 9% (5); in another Brazilian study, CoronaVac booster dose absolute VE was 38% among persons with previous infection (7). CoronaVac booster absolute VE against severe disease during Omicron BA.1/BA.2 period was high in studies in both Hong Kong Special Administrative Region (85%) (4)and Brazil (74%) (5).

The strengths of our study include its prospective cohort design and close follow-up of participants. Moreover, this is one of few studies to evaluate inactivated COVID-19 vaccine in the WHO European Region. Our study also had limitations. The precision of rVE estimates was limited because of the low number of events. We could not measure absolute VE of a COVID-19 booster because of the small number of unvaccinated participants. We were not powered to estimate VE against severe outcomes like hospitalization and death. Most infections occurred during Omicron BA.1/BA.2 circulation, so VE estimates should be understood in that context.

In conclusion, our results highlight the additional preventive benefit of CoronaVac booster vaccine against symptomatic and medically attended infection and provide evidence to encourage uptake of future booster doses among HCWs and the general population in Azerbaijan.

Iris Finci is infectious disease epidemiologist. She is currently a consultant epidemiologist in WHO Regional Office for Europe coordinating COVID-19 and influenza vaccine effectiveness studies in European region. Her primary research interests include viral respiratory infections, tuberculosis and strengthening of disease surveillance.

## Data Availability

The data are not publicly available due to privacy or ethical restrictions. The authors do not have consent from study participants to share the data.

## Author contributions

GH, MAK, NS, SM, and RP conceived the cohort study on which this analysis is based. MAK, LD, NS, SM, CJM, RP, GH, MYRC, and EK planned and implemented the study, including the development of study protocols, data quality checks, and acquisition of data. IF, NS and MAK conceived the article. IF and NS drafted the manuscript and performed the literature search. MYRC performed the data analysis with support from EK. SM, MYRC and EK directly accessed and verified the raw data and take responsibility for the integrity and accuracy of the analyses. BM and CD performed sequencing of study samples. All authors contributed to the interpretation of the results and critically revised the manuscript. All authors had full access to all the data reported in the study and accept responsibility to submit the paper for publication.

## Acknowledgements

Javahir Suleymanova (World Health Organization, Country Office Azerbaijan), Alina Guseinova, Radu Cojocaru (World Health Organization, Regional Office for Europe, Denmark), Trent M. Herdman, Jason Doran (UK Field Epidemiology Training Programme, United Kingdom), Caleb Ward (US Centers for Disease Control, Georgia), Kathryn Lafond (US Centers for Disease Control, USA), Jörn Beheim-Schwarzbach, Victor M. Corman, and Terry C. Jones (Institute of Virology, Charité-Universitätsmedizin Berlin, corporate member of Freie Universität Berlin, Humboldt – Universität zu Berlin, and Berlin Institute of Health, Berlin, Germany; and German Centre for Infection Research (DZIF), partner site Charité, Berlin, Germany); Marta Valenciano and Alain Moren (Epiconcept, France).The Sorgular.az platform was developed by the Public Health and Reform Center of the Azerbaijan Ministry of Health with support from the United Nations Development Program and United Nations Population Fund.

## Disclaimer

The findings and conclusions in this report are those of the author(s) and do not necessarily represent the views of the Centers for Disease Control and Prevention. The authors affiliated with the World Health Organization (WHO) are alone responsible for the views expressed in this publication and they do not necessarily represent the decisions or policies of the WHO.

## Financial statement

This study was funded by The Task Force for Global Health and World Health Organisation Regional Office for Europe.

