## Supplemental materials for "Relative vaccine effectiveness of a CoronaVac booster dose in preventing symptomatic SARS-CoV-2 infection among healthcare workers during an Omicron period in Azerbaijan, January–August 2022"

**Methods – Additional information**

*Enrolment Questionnaire*

Health status of participants was self-assessed by using 5-point scale: excellent, very good, good, fair, poor.

*Case Definition*

Participants who experienced any of the listed symptoms (fever, cough, general weakness, fatigue, headache, muscle ache, sore throat, runny nose, shortness of breath, lack of appetite, nausea, vomiting, diarrhoea, altered mental status, loss of taste, or loss of smell) were tested by PCR for SARS-CoV-2.

*Definition of Prior Infection*

We defined previous infection as a PCR-confirmed infection prior to start of study follow-up, that was documented in either the Etabib database or the MoH/Mandatory Health insurance database.

*Statistical analyses*

In addition to hospital location (urban or sub-urban) and previous SARS-CoV-2 infection that were included in the multivariable model as fixed effects, other potential confounders (e.g., calendar month, age, sex, occupation, hands-on care, household size, chronic condition, smoking, health status) were selected using augmented backward elimination with a change-in-estimate criterion of 5% and a significance threshold of 0.2 (1), however none were retained in the final model.

*Assessment of Time Since Vaccination*

We assessed the effect of time since vaccination on rVE against symptomatic infection over the following time intervals: 7–89, 90–150 and ≥150 days following booster dose. These periods were selected based on empirical information (power to estimate VE) and on bibliography (presence of neutralising antibodies 7 days after receiving booster dose(2), meta-analysis showing that 5 months after booster dose, VE dropped below 20% (3) ). We included symptomatic participants if they had symptoms in the window 14 days before and 4 days after the positive SARS-CoV-2 test.

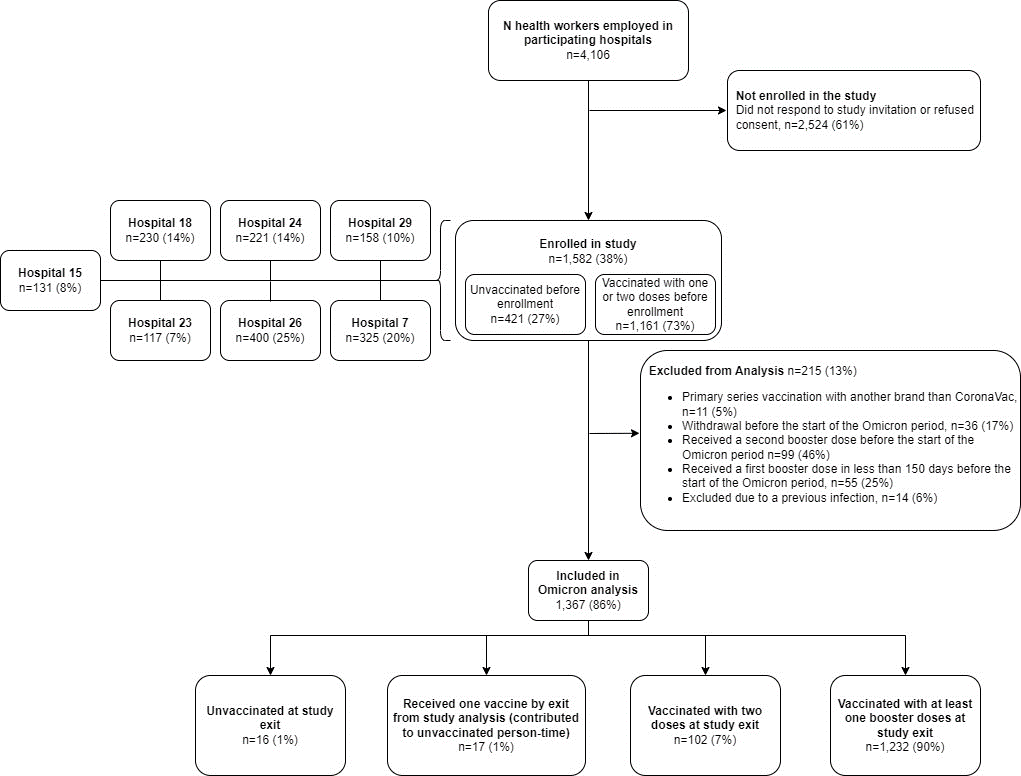

**Figure S1. Flowchart illustrating the enrolment of healthcare workers in COVID-19 vaccine effectiveness study, Azerbaijan, 2022**

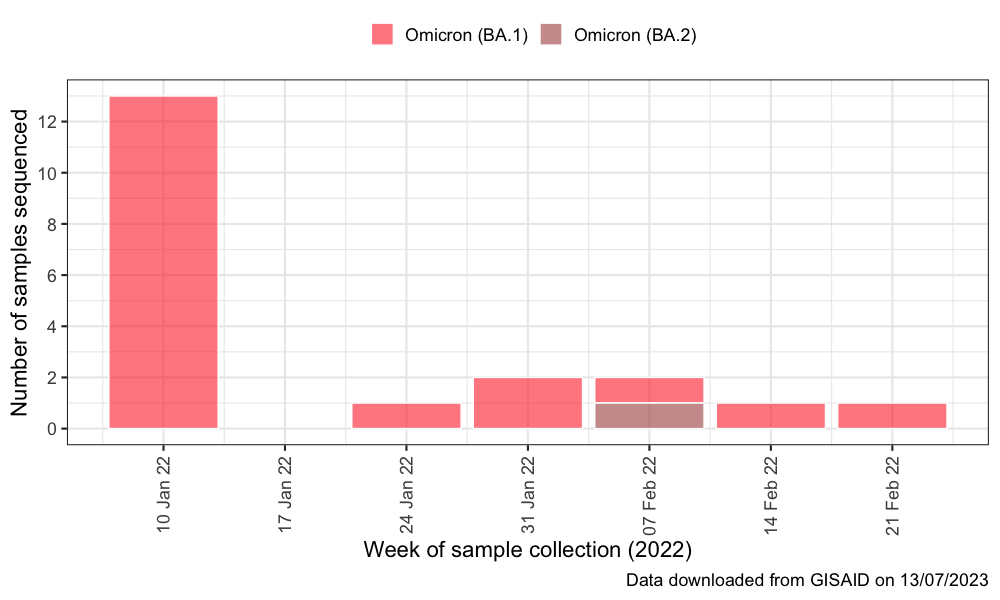

**Figure S2. Whole genome sequencing results from SARS-CoV-2 positive samples from the study and from general surveillance data, GISAID, Azerbaijan, December 2021 – August 2022**

Study samples (N=8), surveillance samples (N=12)

**Table S1. Table 1. Demographic and occupational characteristics of health care worker participants by vaccination status on the last day of the follow-up period (n=115), Azerbaijan, 2022**

|  | **All Participants, N=1367** | **Unvaccinated, N=16** | **Partially vaccinated (one dose), N=17** | **Vaccinated with primary vaccine series (two doses), N=102** | **Vaccinated with booster dose (third dose), N=1232** |
| --- | --- | --- | --- | --- | --- |
| **Age, N=1367** | | | | | |
| Median (IQR) | 48 (39-57) | 46 (33.8-62) | 46 (35-59) | 47.5 (39-58) | 48 (39-57) |
| **Age group, years, N=1367** |  |  |  |  |  |
| 20-29, n (%) | 73 (5) | 3 (19) | 0 (0) | 10 (10) | 60 (5) |
| 30-39, n (%) | 290 (21) | 4 (25) | 7 (41) | 20 (20) | 259 (21) |
| 40-49, n (%) | 371 (27) | 1 (6) | 2 (12) | 24 (24) | 344 (28) |
| 50-59, n (%) | 416 (30) | 3 (19) | 6 (35) | 24 (24) | 383 (31) |
| 60+, n (%) | 217 (16) | 5 (31) | 2 (12) | 24 (24) | 186 (15) |
| **Sex, N=1367** | | | | | |
| M, n (%) | 91 (7) | 1 (6) | 1 (6) | 8 (8) | 81 (7) |
| F, n (%) | 1276 (93) | 15 (94) | 16 (94) | 94 (92) | 1151 (93) |
| **Hospital, N=1367** | | | | | |
| Hospital 15, n (%) | 122 (9) | 0 (0) | 0 (0) | 2 (2) | 120 (10) |
| Hospital 18, n (%) | 204 (15) | 1 (6) | 1 (6) | 8 (8) | 194 (16) |
| Hospital 23, n (%) | 95 (7) | 3 (19) | 2 (12) | 15 (15) | 75 (6) |
| Hospital 24, n (%) | 191 (14) | 3 (19) | 4 (24) | 27 (26) | 157 (13) |
| Hospital 26, n (%) | 359 (26) | 1 (6) | 8 (47) | 28 (27) | 322 (26) |
| Hospital 29, n (%) | 127 (9) | 5 (31) | 1 (6) | 14 (14) | 107 (9) |
| Hospital 7, n (%) | 269 (20) | 3 (19) | 1 (6) | 8 (8) | 257 (21) |
| **Hospital Location, N=1367** | | | | | |
| Urban, n (%) | 517 (38) | 4 (25) | 5 (29) | 37 (36) | 471 (38) |
| Sub-urban, n (%) | 850 (62) | 12 (75) | 12 (71) | 65 (64) | 761 (62) |
| **Occupation/Role in hospital, N=1367** | | | | | |
| Nurse or Midwife, n (%) | 506 (37) | 4 (25) | 6 (35) | 36 (35) | 460 (37) |
| Medical Doctor, n (%) | 349 (26) | 5 (31) | 6 (35) | 23 (23) | 315 (26) |
| Other, n (%) | 512 (37) | 7 (44) | 5 (29) | 43 (42) | 457 (37) |
| **BMI, N=1367** | | | | | |
| Underweight or normal, n (%) | 382 (28) | 7 (44) | 3 (18) | 24 (24) | 348 (28) |
| Overweight, n (%) | 509 (37) | 6 (38) | 5 (29) | 37 (36) | 461 (37) |
| Obese, n (%) | 476 (35) | 3 (19) | 9 (53) | 41 (40) | 423 (34) |
| **Smoking, N=1367** | | | | | |
| Currently smokes, n (%) | 38 (3) | 1 (6) | 0 (0) | 1 (<1) | 36 (3) |
| Never smokes, n (%) | 1315 (96) | 15 (94) | 15 (88) | 101 (>99) | 1184 (96) |
| Smoked previously, n (%) | 14 (1) | 0 (0) | 2 (12) | 0 (0) | 12 (<1) |
| **Any chronic condition, N=1367** | | | | | |
| No, n (%) | 820 (60) | 10 (62) | 9 (53) | 53 (52) | 748 (61) |
| Yes, n (%) | 547 (40) | 6 (38) | 8 (47) | 49 (48) | 484 (39) |
| **Number of chronic conditions, N=1367** | | | | | |
| 0, n (%) | 820 (60) | 10 (62) | 9 (53) | 53 (52) | 748 (61) |
| 1, n (%) | 383 (28) | 4 (25) | 4 (24) | 27 (26) | 348 (28) |
| ≥2, n (%) | 164 (12) | 2 (12) | 4 (24) | 22 (22) | 136 (11) |
| **Number of household members, N=1280** | | | | | |
| 1-3, n (%) | 500 (39) | 9 (60) | 9 (53) | 32 (34) | 450 (39) |
| 4-5, n (%) | 654 (51) | 3 (20) | 7 (41) | 51 (54) | 593 (51) |
| ≥6, n (%) | 126 (10) | 3 (20) | 1 (6) | 11 (12) | 111 (10) |
| **Hands on care, N=1367** | | | | | |
| No, n (%) | 798 (58) | 10 (62) | 11 (65) | 62 (61) | 715 (58) |
| Yes, n (%) | 569 (42) | 6 (38) | 6 (35) | 40 (39) | 517 (42) |
| **Delay between 2nd dose and end of follow-up (days), N=1334** | | | | | |
| Median (IQR) | 328 (219-346) | _ | _ | 185 (165-241) | 333 (282.2-346) |
| **Delay between 3rd dose and end of follow-up (days), N=1232** | | | | | |
| Median (IQR) | 94 (55.2-117.5) | _ | _ | _ | 94 (55.2-117.5) |
| **COVID-19 vaccination status and vaccine brand at study period start, N=1367** | | | | | |
| Unvaccinated, n (%) | 16 (1) | 16 (100) |  |  |  |
| CoronaVac - 1 dose, n (%) | 17 (1) |  | 17 (100) |  |  |
| CoronaVac - 2 doses, n (%) | 102 (7) |  |  | 102 (100) |  |
| CoronaVac - 3 doses, n (%) | 1186 (87) |  |  |  | 1186 (96) |
| 2 doses CoronaVac + 1 dose BNT162b2, n (%) | 46 (3) |  |  |  | 46 (4) |

Table S2. Table with GISAID sample IDs and information about the authors

(website: https://epicov.org/epi3/epi_set/230713gx?main=true)

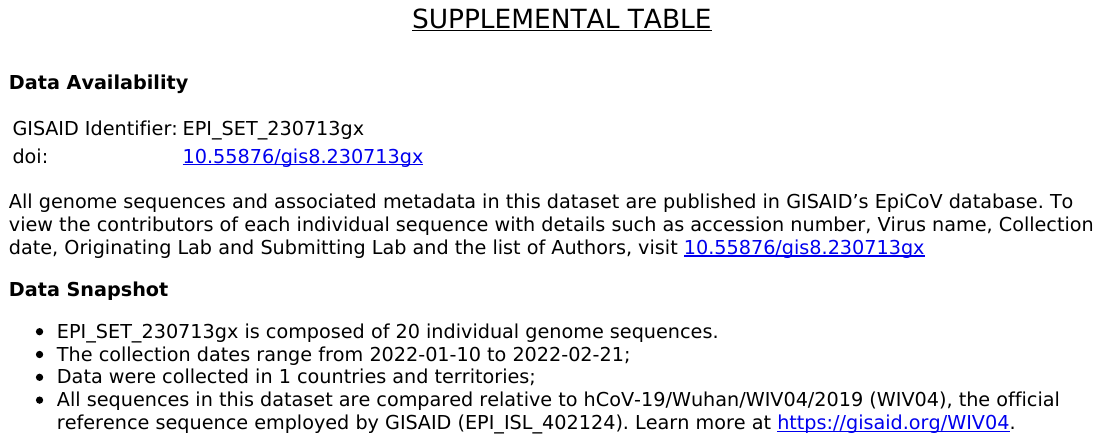
